# Sphingosine-1-phosphate – sphinganine-1-phosphate Imbalance Drives Airway Hyperreactivity

**DOI:** 10.64898/2026.03.15.26348448

**Authors:** Andrea F. Heras, Sarah Brown, Tilla S. Worgall, Jose F. Perez-Zoghbi, Stefan Worgall

## Abstract

Asthma is the most common chronic respiratory disease of childhood and is strongly associated with genetic variants at the 17q21 locus that increase expression of ORMDL3, a negative regulator of serine palmitoyl-CoA transferase (SPT), the rate-limiting enzyme in de novo sphingolipid synthesis. Reduced sphingolipid production has been linked to airway hyperreactivity, a key physiological feature of asthma, but the mechanisms connecting altered sphingolipid metabolism to airway dysfunction remain unclear.

We examined whether sphingolipid metabolites regulate airway smooth muscle reactivity. Circulating sphingolipids were quantified in children with asthma carrying 17q21 risk alleles and in mice with reduced SPT activity. Functional airway responses were assessed in precision-cut lung slices exposed to sphingosine-1-phosphate (S1P), sphinganine-1-phosphate (Sa1P), and S1P receptor antagonists. Homozygous carriers of the rs7216389 risk allele and SPT-deficient mice displayed an increased S1P-to-Sa1P ratio. In functional assays, Sa1P opposed S1P-induced airway contraction, and increasing Sa1P availability reduced airway hyperresponsiveness. These findings identify the S1P/Sa1P axis as a metabolic rheostat regulating airway smooth muscle tone and suggest that targeting sphingolipid metabolism may offer a therapeutic strategy to mitigate intrinsic airway hyperreactivity in asthma.

**One sentence summary:** An imbalance between sphingosine-1-phosphate and sphinganine-1-phosphate links the asthma risk locus 17q21 to airway hyperreactivity and reveals sphingolipid metabolism as a potential therapeutic target.

## Introduction

Asthma is a chronic respiratory condition characterized by airway inflammation and hyperreactivity. Genetic predisposition, particularly the 17q21 locus, plays a crucial role in the risk of childhood asthma. (*1–3*) Among the implicated genes, gasdermin B (*GSDMB*) and Oromucoid-like 3 (*ORMDL3*) have distinct functions(*4–7*): *GSDMB* modulates inflammation through pyroptosis,(*3*) whereas *ORMDL3* regulates de novo sphingolipid synthesis via serine palmitoyl-CoA transferase (SPT).(*8–10*) Rs7216389 is a single-nucleotide 17q21 polymorphism (SNP) is among the strongest associations in childhood-onset asthma (Moffatt 2007) and the asthma risk allele (T) increases expression of *ORMDL3*, especially in blood and airway smooth muscle.(*1, 11–15*) Altered *ORMDL3* expression can lead to dysregulated sphingolipid metabolism.(*16*) Despite its association with airway hyperreactivity,(*13, 17*) the precise role of *ORMDL3*-dependant sphingolipid alterations in asthma pathophysiology remains unclear.

*ORMDL3*-mediated inhibition of SPT reduces de novo sphingolipid synthesis, leading to decreased sphinganine levels, its derivative, Sa1P, and downstream sphingolipids (**Supplemental Figure 1**).(*8, 10*) These sphingolipids are critical for maintaining cellular homeostasis, yet their impact on airway reactivity remains underexplored. Previous studies have demonstrated that children with asthma and 17q21 risk alleles exhibit reduced sphingolipid synthesis, which correlates with increased airway reactivity.(*13, 18, 19*) Furthermore, SPT-deficient mice display airway hyperresponsiveness without concurrent inflammation(*20*), (*17*) suggesting a direct link between sphingolipid metabolism and airway smooth muscle contractility.

Metabolomic studies highlight the role of sphingolipid metabolism in asthma.(*21–23*) Asthmatic children show sphingolipid variations linked to atopy and inflammation, likely affecting ceramide and sphingolipid levels.(*21, 24, 25*) While asthma-related inflammation is thought to increase S1P levels,(*26–29*) reduced sphingolipid synthesis does not. However, *ORMDL3* knockdown in airway epithelium elevates S1P levels.(*30*) S1P is a key sphingolipid mediator^24^ known to induce airway smooth muscle contraction via S1P receptors 2 and 3.^21,25^ However, the role of Sa1P, a structurally similar but functionally distinct metabolite, remains unknown. Given the structural similarities between S1P and Sa1P,(*31–33*) we hypothesized that Sa1P counteracts S1P-mediated airway contraction. To investigate this, we examined blood sphingolipid profiles in children with asthma-risk alleles and SPT-deficient mice and assessed airway responses to S1P and Sa1P in precision-cut lung slices (PCLS).

This study reveals that 17q21 asthma-risk alleles, associated with decreased sphingolipid de novo synthesis and SPT deficiency, lead to an imbalance between S1P and Sa1P, resulting in increased small airway reactivity. Restoring this imbalance by augmenting Sa1P in the small airways reduces airway hyperreactivity. These findings support sphingolipid synthesis as a potential therapeutic target in asthma.

## Results

### Asthma-associated 17q21 risk allele and SPT-deficiency increase the ratio S1P to Sa1P

To evaluate whether dysregulation of sphingolipid metabolism in asthma modulates airway reactivity,(*17*) we quantified S1P (**Figure 1a**) and Sa1P (**Figure 1b**) in the blood of children with asthma-associated genotype for the common 17q21 SNP rs7216389 (T and C alleles), with and without asthma.(*17*) Among the 342 participants, the TT genotype was more prevalent in children with asthma compared to controls (63% vs. 33%, p < 0.001). No differences were observed in asthma severity distribution among the asthma group (p > 0.9). Additionally, there were no significant differences between groups in terms of sex (p = 0.2) or age (p = 0.5), but a difference was noted on race with a higher proportion of White children and a lower proportion of Black children in the control group (p < 0.001), Ethnicity also showed a modest difference (p = 0.033), with a greater proportion of Hispanic or Latino children in the asthma group. (**Supplemental Table 1.**)

**Figure 1.**
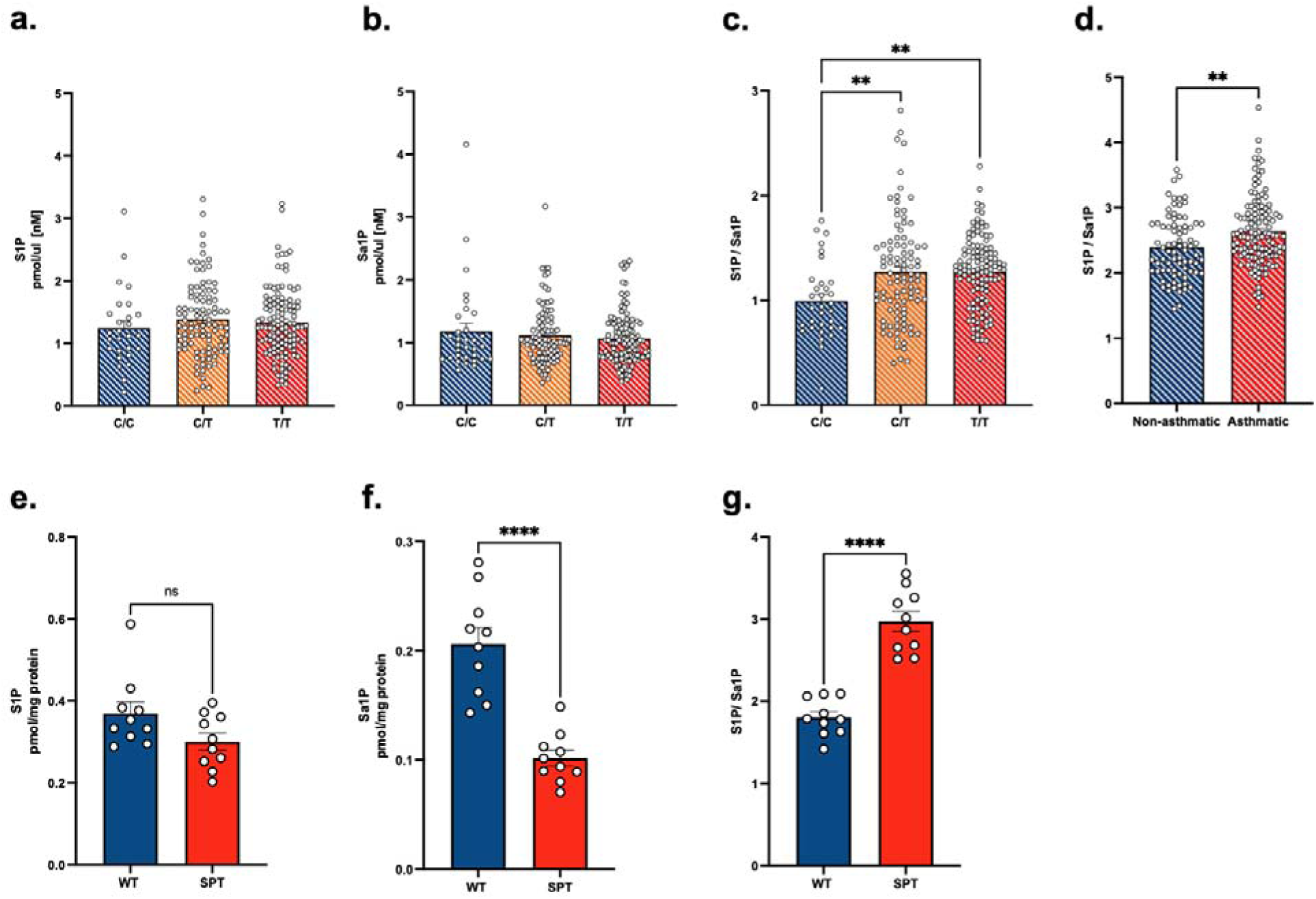
The blood S1P/Sa1P ratio is increased in the presence of the rs72316389 asthma-risk allele and asthma in children, and in mice with impaired sphingolipid synthesis. Sphingosine-1-phosphate (S1P) **(a),** Sphinganine-1-phosphate (Sa1P) **(b),** and S1P to Sa1P **(c)** concentration ratios (pM/pM) in whole blood of children grouped by the presence of rs7216389 T (asthma-risk) and C (non-risk) alleles, or **(d)** asthma; and **(e, f and g)** wild-type Sptlc2^+/+^ (WT) and SPT-deficient Sptlc2^+/−^ (SPT) mice. The presence of T alleles (CT or TT), asthma, and SPT-deficiency is associated with a higher S1P/Sa1P ratio. \**p* < 0.05, one-way ANOVA with multiple comparisons test; **p< 0.01, unpaired t-test.

Despite no significant variation in individual metabolites, the post hoc analysis revealed that the ratio of S1P to Sa1P ratio is higher in children who were homozygous for the asthma risk T allele compared to those who were homozygous for the non-risk C allele (F 2, 199 = 3.326, P = 0.038, η^2^=0.032) (**Figure 1c**) and in children with asthma compared to those without asthma (p= 0.001, df=203) (**Figure 1d**). Furthermore, in an animal model with impaired *de novo* sphingolipid synthesis, which has increased airway reactivity, Sa1P levels were predictably reduced (p=0.001, df= 8) relative to S1P (**Figure 1e, f**). Similarly to children, the S1P/Sa1P ratio was increased in SPT-deficient (Sptlc2^+/−^) mice compared to their wild-type (WT, Sptlc2^+/+^) littermates (p= 0.001, df= 14) (**Figure 1g**). These findings suggest that asthma risk alleles affecting sphingolipid synthesis, as well as the diagnosis of asthma, are associated with an increased S1P/Sa1P ratio and that the SPT-deficient mice is a good model to study sphingolipid-associated airway hyperreactivity.

### S1P, but not Sa1P, induces peripheral airway contraction

To determine whether an imbalance in S1P and Sa1P contributes to airway hyperresponsiveness, we investigated their effects on airway smooth muscle contraction. While S1P is known to activate airway smooth muscle contraction,(*34*) the effects of Sa1P remain unknown. We characterized the contractile responses of peripheral airways to S1P and Sa1P using PCLS from SPT-deficient and wildtype (WT) mice (p= 0.001) (**Figure 2**). In SPT-deficient mice, S1P induced a concentration-dependent contraction of the peripheral airways, with a maximal lumen reduction of 34.2 ± 12.7% and a stimulatory concentration 50 (SC_50_) of 1 µM. In contrast, Sa1P had a minimal effect, with a maximal contraction of only 6.6 ± 10.3 % (**Figures 2a-c**). Notably, we observed that S1P-induced airway contraction ceased upon S1P withdrawal, demonstrating the response’s reversibility and suggesting a receptor-mediated mechanism in airway smooth muscle.

**Figure 2.**
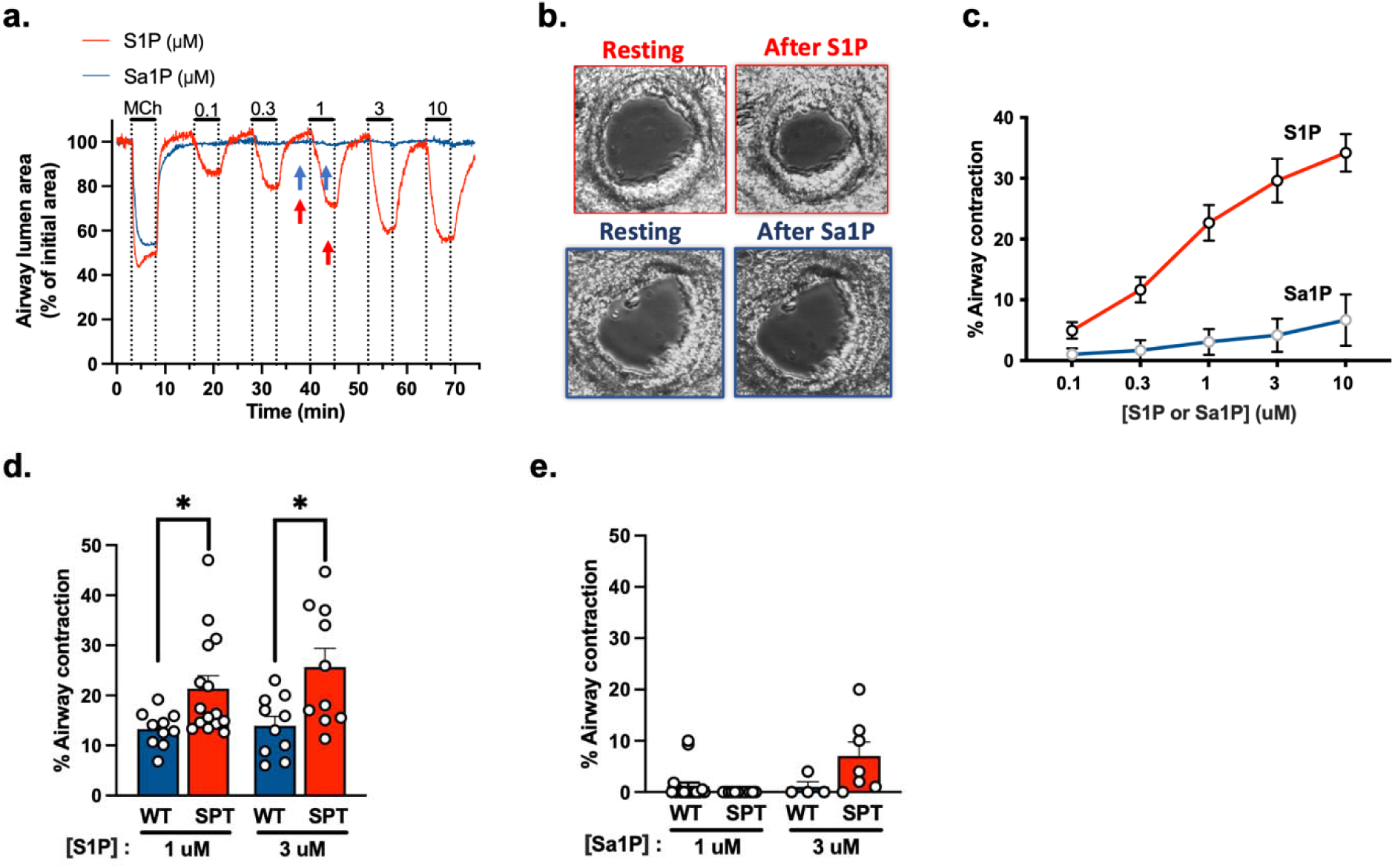
Peripheral airway contractile responses to S1P and Sa1P. Peripheral airway contractility in response to S1P, Sa1P, and methacholine (MCh) was assessed in precision-cut lung slices (PCLS) from SPT-deficient (SPT) or wild-type mice (WT). (**a)** Representative traces of the cross-sectional airway luminal area changes over time in response to MCh (0.3 µM) and subsequent exposure to increasing concentrations of S1P and Sa1P, as indicated above the traces. **(b)** Phase-contrast microscope images of two representative PCLS from SPT-deficient mice showing peripheral airways before and after exposure to either S1P or Sa1P (3 µM each) and recorded at the times indicated by the arrows in the corresponding traces in **b**. (**c)** Concentration-dependent peripheral airway contraction to S1P and Sa1P. Data are means ± SEMs of 18–24 airways from 6 mice. (**d, e)** Peripheral airway contraction in PCLS from WT and SPT mice in response to either (d) S1P or (e) Sa1P (each 1 and 3 _μ_M). Data are means ± SEMs of 6–19 airways. \**p* < 0.05, unpaired *t*-test.

Next, we compared the effects of S1P and Sa1P in PCLS from SPT-deficient and WT mice at two S1P concentrations (1 and 3 μM Figures 2d and **e**). Airways from WT mice exhibited a weaker yet significant contractile response to S1P compared to those from SPT-deficient mice (1μM p=0.025, df=23 and 3μM p=0.012, df=18) (**Figure 2d**). However, Sa1P did not induce significant contraction in airways from either SPT-deficient or WT mice (1μM p=0.198, df=29 and 3μM p=0.150, df=9) (**Figure 2e**). These findings demonstrate that S1P induces a rapid and reversible contraction of the peripheral airways with a more pronounced effect in SPT-deficient mice, whereas Sa1P has little to no contractile effect.

### Ca^2+^ Oscillations in airway smooth muscle cells induced by S1P and Sa1P

To determine whether S1P, but not Sa1P, stimulates airway smooth muscle cells (SMCs) to induce airway contraction, we examined changes in intracellular Ca² in response to S1P and Sa1P and compared them to the response elicited by methacholine (MCh). For these studies, we prepared PCLS from WT (Sptlc2^+/+^) mice expressing the intracellular calcium reporter GCAMP6 and used a custom-built video-rate confocal microscope to monitor intracellular Ca^2+^ dynamics in peripheral airway SMCs (**Figure 3a**). First, PCLS were exposed to 0.3 µM MCh, and intracellular Ca^2+^ signaling was recorded for two minutes. The MCh was then washed out with sHBSS followed by stimulation with either S1P or Sa1P (3 µM each). S1P stimulation triggered a sustained increase in intracellular calcium concentration [Ca^2+^]_i_ characterized by persistent Ca^2+^ oscillations similar to those induced by MCh, though with a slightly slower frequency. Concurrent airway lumen measurements revealed that airways relaxed upon MCh washout (increased luminal area) and contracted again as Ca^2+^ oscillations resumed in response to S1P (**Figure 3b**); conversely, the addition of Sa1P resulted in no Ca^2+^ oscillations and no airway contraction (**Figure 3c**). The mean Ca^2+^ oscillation frequency averaged 15 ± 3 oscillation/min for MCh and 12 ± 3 oscillation/min for S1P (F 2, 22 = 7.583, P = 0.003, η^2^=0.408) (**Figure 3d**). These findings suggest that S1P receptors may be coupled to intracellular signaling pathways similar to those of muscarinic receptors, which mediate MCh-induced airway contraction.

**Fig 3.**
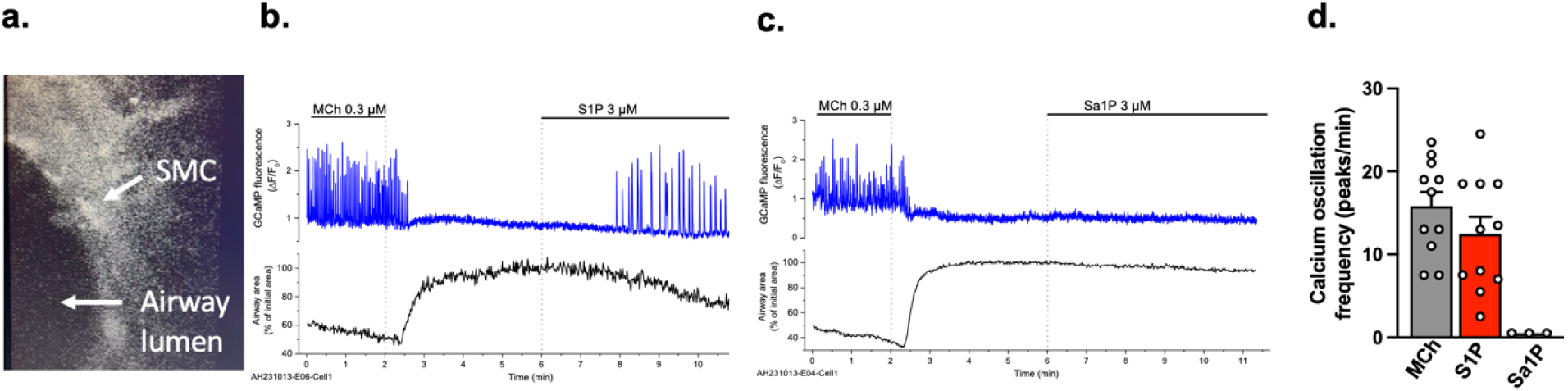
[Ca^2+^]_i_ signaling is induced by S1P, but not by Sa1P, in airway smooth muscle cells. Intracellular Ca^2+^ oscillations were assessed by confocal microscopy in single-airway smooth muscle cells in PCLCs from WT mice expressing the intracellular Ca^2+^ reporter GCaMP6. (**a).** Representative fluorescence image showing part of a peripheral airway depicting the airway lumen and an airway smooth muscle cell (SMC). (**b, c).** Representative traces of Ca^2+^ oscillations in SMC (blue line) and airway lumen (black line) following stimulation with MCh (0.3 µM) and subsequently with either (b) S1P or (c) Sa1P (each 3 µM). (**d).** Summary of the frequency of Ca^2+^ oscillations stimulated by MCh, S1P, and Sa1P in experiments like those shown in b and c. Data means ± SEMs of 5 to 8 airways from 4 mice.

### Supplementing Sa1P decreases peripheral airway contractility to S1P and MCh

Given our findings that the S1P/Sa1P ratios are elevated in children carrying the asthma-risk T allele and in SPT-deficient mice, we sought to elucidate the mechanism linking altered sphingolipid metabolism to airway reactivity. We first hypothesize that Sa1P attenuates the contractile response of peripheral airways to S1P. To test this, PCLS from SPT mice were exposed to S1P (1 µM) alone or in combination with increasing concentrations of Sa1P (0.5 and 1 µM). As hypothesized, the results demonstrated that peripheral airway contraction decreased with increasing Sa1P concentrations, supporting the idea that Sa1P plays a crucial role in counteracting S1P-induced airway contraction (F 2, 90 = 4.970, P = 0.009, η^2^=0.099) (Figures 4a and **b**).

**Fig 4.**
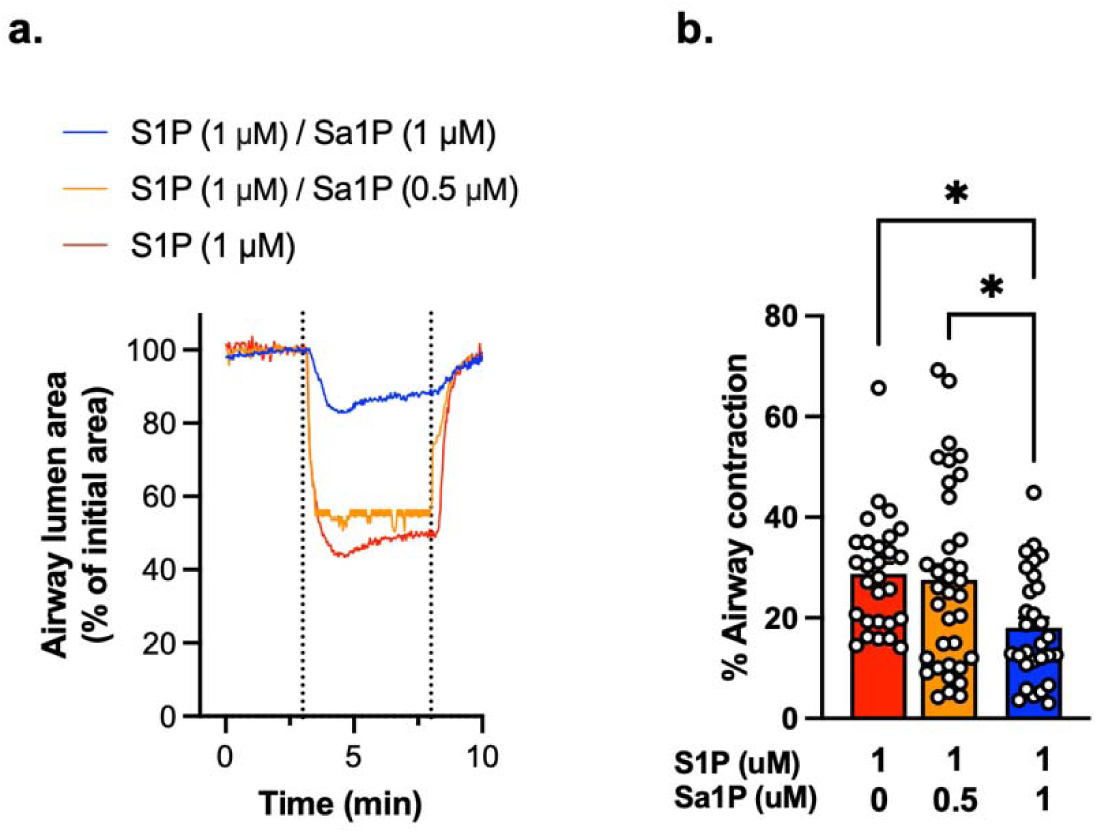
S1P-induced airway contraction is decreased in the presence of increasing Sa1P concentrations. **(a)**. Representative experiment showing the contractile responses of peripheral airways to decreased S1P/Sa1P ratio concentrations in PCLS from WT mice. (**b).** Summary of the airway contraction stimulated by different S1P/Sa1P ratios showing decreased contraction at a ratio 1:1. Contraction was calculated from experiments like those illustrated in A, and data are means ± SEMs of 32-48 PCLS from six mice in each group, \**p* < 0.05, Kruskal-Wallis test.

Next, we investigated whether chronic Sa1P exposure could reduce the hypercontractility of SPT-deficient airways to MCh. We previously reported that SPT-deficient mice exhibit enhanced MCh-induced airway contraction.(*17*) Here, we hypothesized that supplementing Sa1P (but not S1P) would restore normal airway contractility, given that Sa1P levels are reduced in SPT-deficient mice. To test this, we incubated PCLS from SPT-deficient mice for 15 hours with Sa1P, S1P, Sa1P + S1P, or DMSO (control) before assessing MCh-induced contraction. Notably, Sa1P alone or in combination with S1P reduced MCh-induced airway contraction compared to S1P alone and the DMSO control (F 3, 80 = 2.7, P = 0.048, η^2^=0.093) (Figures 5a and **b**). These findings suggest that restoring Sa1P levels in SPT-deficient tissues reverses peripheral airway hyperreactivity, highlighting the crucial role of Sa1P deficiency in airway hyperresponsiveness. Furthermore, the reduced Sa1P levels associated with 17q21 risk alleles affecting *ORMDL3* expression may contribute to airway hyperresponsiveness in these individuals.

**Fig 5.**
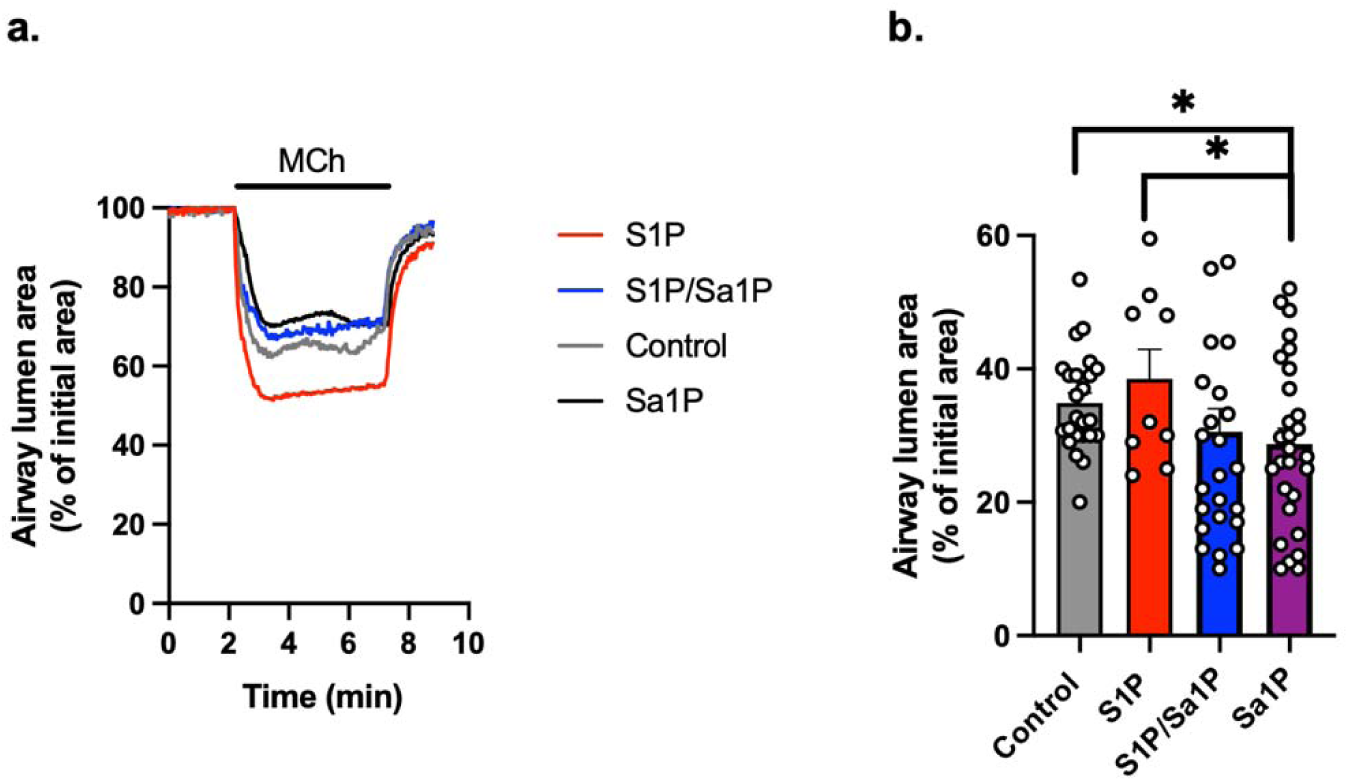
Pre-incubation of PCLS from SPT-deficient mice with Sa1P decreases peripheral airway contractility to muscarinic agonist MCh. **(a)**. Representative traces showing the contractile responses of airways to 0.3 _μ_M MCh and its subsequent washout in PCLS that were incubated with S1P 1 _μ_M, S1P 1 _μ_M/Sa1P 1 _μ_M, Sa1P 1 _μ_M or vehicle (control) for 15 hours in cell culture medium at 37°C. (**b).** Summary data showing the effect of the PCLS overnight incubation with the sphingolipids on the MCh-induced airway contraction obtained from experiments like those illustrated in **a**. Data are means ± SEMs of **28–32** airways in different PCLS from four SPT mice in each group. \**p* < 0.05. Kruskal-Wallis test.

### S1P receptor 2 antagonist inhibits S1P-induced peripheral airway contraction

To identify the S1P receptor mediating peripheral airway contraction, we tested two specific S1P receptor antagonists targeting the S1P G-protein coupled receptors 2 (S1PR2) (**Figures 6a**) and 3 (S1PR3) (**Figures 6d)**. PCLS from WT mice were exposed to S1P and Sa1P before and after incubation with either S1PR2 antagonist JTE-013 or S1PR3 antagonist CAY10444. Our results demonstrate that S1PR2 inhibition reduced S1P-induced airway contraction by approximately 55% (**Figures 6b** p=0.001, df=10 and **c** p=0.023, df=6), whereas S1PR3 inhibition had no significant effect (**Figures 6e** p=0.541, df=6 and **f** p=0.870, df=6). No changes in airway contractility were observed with Sa1P alone (**Supplemental Fig. 2**). These findings suggest that S1PR2 is a crucial mediator in S1P-induced airway contraction, whereas S1PR3 seems to have a minimal role in this response. Furthermore, Sa1P alone has no contractile effect on S1P receptors.

**Fig 6.**
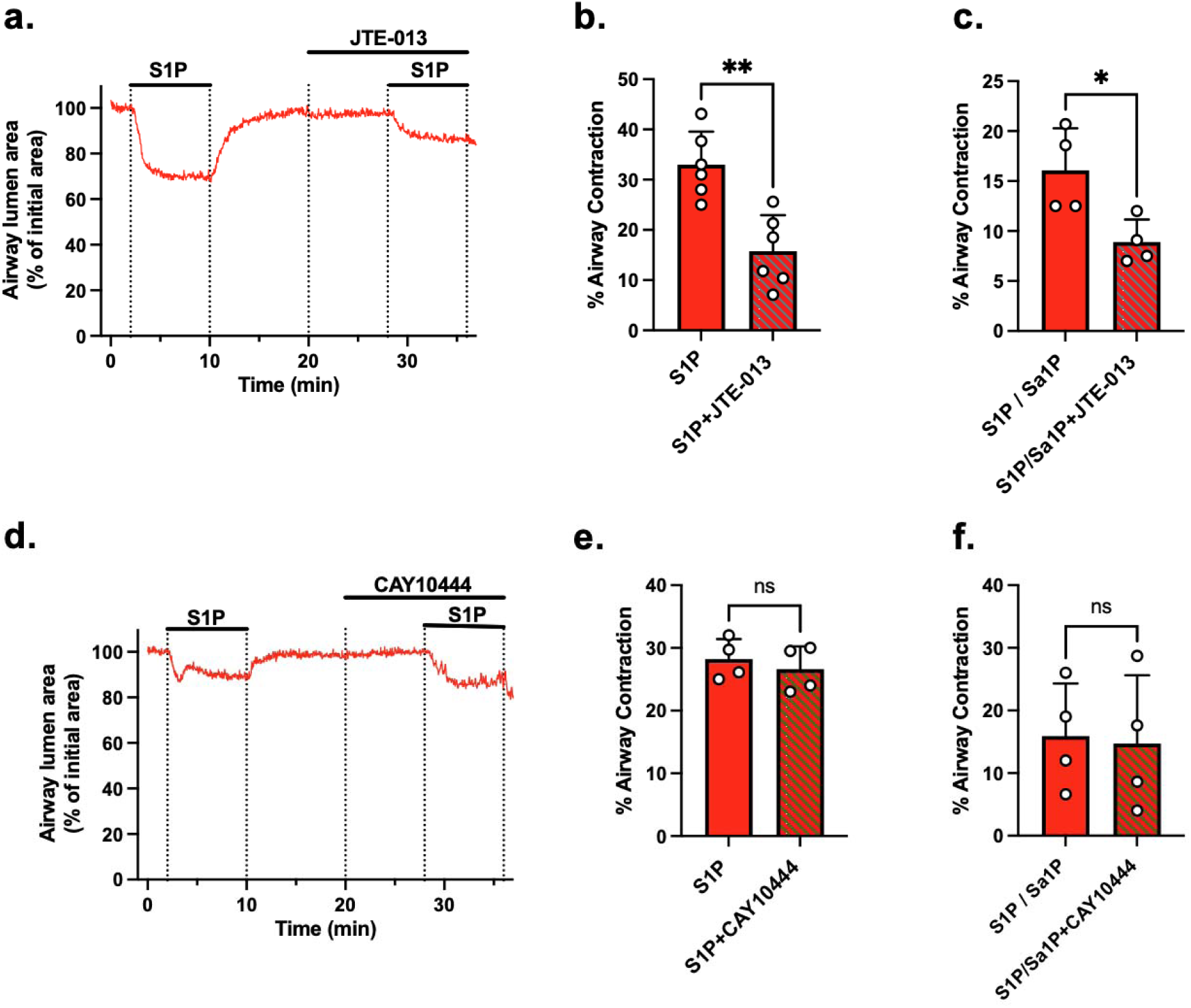
The effect of S1P receptor antagonists on airway contraction stimulated by S1P. (**a, d)** Representative traces showing airway contraction induced by S1P (1 µM) before and after pre-exposure of the PCLS to S1P receptor 2 antagonist JTE-013 (10 µM) and S1P receptor 3 antagonist CAY10444 (10 µM). (**b, c, e, f**) Summary data showing the effect of the inhibitors on the S1P (**b** and **e**) and S1P/Sa1P (**c** and **f**)-induced airway contraction. The contraction was calculated from experiments like those illustrated in **a** and **d**, and data are means ± SEMs of 5–10 airways from four mice in each group. \*\**p*< 0.01, unpaired *t*-test.

## Discussion

A link between asthma and altered sphingolipid metabolism has been established, particularly involving plasma sphingolipids like ceramides. In conditions such as asthma, atopy, and other inflammatory disorders, plasma sphingolipids, especially ceramides, are predominantly elevated.(*19, 21, 22, 24, 25*) Conversely, *de novo* sphingolipid synthesis is reduced in the blood cells of children carrying 17q21 asthma-risk genotypes, which increases *ORMDL3* expression.(*13, 18*) Our study builds on these findings by showing that the S1P/Sa1P ratio is higher in the blood of children with asthma and those carrying the asthma-risk rs7216389 T allele compared to non-asthmatic children and those without the risk allele. Additionally, we observed a similar elevation of the S1P/Sa1P ratio in Sptlc2 heterozygous knock-out mice, a murine model of reduced *de novo* sphingolipid synthesis in asthma.(*20*) This suggests that Sa1P deficiency, resulting from impaired de novo sphingolipid synthesis, could shift the balance towards S1P-driven pathways, potentially contributing to airway hyperreactivity.

S1P is known to modulate airway smooth muscle contraction and bronchial hyperresponsiveness.(*28, 34, 35*) However, the precise mechanisms remain debated. Maguire et al. challenged this concept by reporting that S1P does not induce direct airway contraction in human or murine bronchioles and does not elicit bronchoconstriction upon inhalation in humans.(*28*) However, they observed that S1P enhances methacholine-induced airway reactivity, suggesting an indirect role. In contrast, our data provide direct evidence that S1P induces dose-dependent contraction of small airways in PCLS. These discrepancies may arise from methodological differences, including PCLS vs. wire myography, airway size variations, and the solubilization method of S1P (see Methods), which can influence S1P bioavailability in the experimental solutions.

Additionally, our direct visualization of Ca^2+^ oscillations in airway SMCs supports the idea that S1P functions as a bronchoconstrictor by directly stimulating ASMCs in this ex vivo tissue preparation. The similarity of the Ca^2+^ signaling induced by S1P and MCh suggests a common stimulus-contraction coupling pathway downstream of the S1PR and muscarinic receptors. We previously demonstrated that ACh stimulation of muscarinic receptors is associated with activation of phospholipase C and inositol triphosphate (IP_3_) induced Ca^2+^ release from internal stores in the peripheral airways.(*36*) We propose a similar mechanism coupling the activation of S1P receptors with the ASMC contraction in the peripheral airways.

A novel finding of our study is that Sa1P does not induce airway contraction, unlike S1P, despite their structural similarity. This implies that Sa1P either binds to S1P receptors but cannot stimulate them or lacks a receptor system. Notably, supplementing Sa1P dose-dependently reduced S1P-induced airway contraction, supporting a potential antagonistic role of Sa1P against S1P-mediated hyperreactivity. We also showed that restoring Sa1P levels in SPT-deficient airways mitigated hyperreactivity to methacholine, underscoring that Sa1P’s ability to counterbalance S1P-driven deficiency is associated with airway hyperresponsiveness. This effect may arise from restoring S1P/Sa1P balance, thereby normalizing sphingolipid signaling within ASMCs. Our findings suggest that Sa1P plays a protective role against airway hypercontractility, offering a potential therapeutic avenue for conditions where sphingolipid dysregulation exacerbates asthma symptoms.

To identify the receptor mediating S1P-induced airway contraction, we selectively inhibited S1PR2 and S1PR3. Our data indicate that S1PR2 blockade significantly reduced S1P-induced contraction, whereas S1PR3 inhibition had no effect. This suggests that S1PR2 may play a key role in mediating S1P-driven airway hyperreactivity and that targeting S1PR2 could be a possible approach for modulating S1P-mediated bronchoconstriction.

We recognize that SPT-deficient mice do not fully recapitulate the complex genetic and environmental factors influencing asthma in humans, particularly individuals with the 17q21 risk allele. While the murine model effectively mimics reduced *de novo* sphingolipid synthesis, future studies should validate these findings in human-derived airway tissues or patient cohorts. Although PCLS-based airway contraction assays are the best available *in situ* preparation to study peripheral airway reactivity to bronchoconstrictors, as they preserve most of the multicellular interactions present in the lung microenvironment, the possible effects of sphingolipid synthesis in epithelial cells, the release of immune mediators, and the extracellular matrix components were not evaluated in this work. In addition, sphingolipid-driven airway remodeling was not addressed in this study. Further exploration using organoid models, co-culture systems, or in vivo asthma models could enhance translational relevance.

Our findings reveal a functional consequence of an imbalanced S1P/Sa1P ratio in airway hyperreactivity, with direct implications for asthma risk in individuals carrying the 17q21 risk allele. Targeting the S1P/Sa1P balance or selectively inhibiting S1PR2 could offer novel therapeutic strategies to mitigate airway hyperresponsiveness. Future investigations should explore: (1) the mechanistic underpinnings of Sa1P’s inhibitory effect on S1P-induced airway contraction, (2) the potential of Sa1P analogs or S1PR2-specific inhibitors in asthma therapy, and (3) the role of S1P/Sa1P balance in other inflammatory airway diseases.

In conclusion, our study establishes Sa1P as a functional antagonist of S1P in airway smooth muscle, with implications for airway hyperreactivity linked to impaired sphingolipid synthesis. By elucidating S1P’s role in airway contraction and the protective function of Sa1P, we provide new insights into sphingolipid-mediated airway physiology and potential therapeutic targets for asthma.

## Materials and Methods

This study accounted for sex as a biological variable. For the human cohort, both males and females were included to ensure generalizability of findings across sexes. In animal experiments, only females were used because sphingolipid levels with SPT deficiency are more pronounced and consistent in female mice, reducing variability and improving reproducibility. Although only one sex was studied in the animal model, the underlying biological mechanisms are expected to be relevant to both sexes. This approach was chosen to balance scientific rigor with feasibility.

### Blood from children with asthma

Children aged 2 to 21 years (n = 242) with and without physician-diagnosed persistent asthma were recruited from pediatric clinics in New York between 2020 and 2025. Written informed consent was obtained from all participants before their inclusion in the study, and written assent was obtained from participants aged 1-21 years. Blood samples were collected for sphingolipid quantification and 17q21 SNP genotyping.

### Sphingolipid analysis

S1P and Sa1P were quantified by high-pressure liquid chromatography-electrospray ionization tandem mass spectrometry (HPLC-MS/MS), with minor modifications to a previously described method (see Supplemental Methods for complete details).(*37*)

### SNP genotyping

Genotyping for the 17q21 single nucleotide polymorphism rs7216389 was performed from blood cell DNA using the TaqMan® SNP Genotyping Assay (SNP ID: rs7216389) in a QuantStudio 6 Flex Real-Time PCR System and analyzed using QuantStudio Software (Applied Biosystems).

### Reagents

S1P and Sa1P were purchased from Avanti Research™ - Polar lipids. S1P and Sa1P were solubilized in ethanol or DMSO, respectively, to prepare 2-mM and 3mM stocks and diluted in Hank’s balanced salt solution (HBSS) or physiological medium to their final concentration on the same day the experiments were performed. JTE-013, an S1P receptor two antagonist (S1P_2_), and CAY10444 S1P receptor three antagonists (S1P_3_) were solubilized in DMSO to prepare 10-mM stocks and diluted in HBSS to their final concentration.

The doses used were chosen based on plasma S1P levels in humans, which are typically in the low nanomolar range, ranging from 200 to 1000 nM (0.2 to 1 µM)(*38, 39*) micromolar concentrations are used to activate S1P receptors in vitro.(*40*) Concentrations in inflamed or cancerous tissues may be higher, but are likely to be below 10 µM.(*27, 41*)

### Mice

SPT-deficient mice (Sptlc2^+/−^) and wild-type (Sptlc2^+/+^) littermates were used to model impaired *de novo* sphingolipid synthesis. All mice were used at 10 to 14 weeks of age. Sptlc2^+/-^mice have approximately 60% decreased SPT activity and lower lung SL, have demonstrated chronic impaired pulmonary SL synthesis and airway hyperreactivity.(*20, 42*)

### Precision-cut lung slices

Precision-cut lung slices (PCLS) were prepared from murine lungs and exposed to S1P, Sa1P, or S1P receptor antagonists. The methods used to prepare PCLS have been described in detail in our prior publication(*17*), and only a brief outline is provided here. PLCS were prepared from 10 to 14-week-old heterozygous SPT-deficient mice (Sptlc2^+/−^) or homozygous wild type (WT) mice (Sptlc2^+/+^). The trachea was cannulated, and the lungs were inflated with 1.2 ± 0.1 ml of 2% agarose (low-gelling temperature) in Hanks’ balanced salt solution supplemented with 20 mM HEPES pH=7.40 (sHBSS), followed by 0.2 ml of air to flush the agarose out of the airways and into the distal alveolar space. Each lung lobe was removed and cut into serial sections of 130-µm thick with a vibratome at 4°C. Slices were maintained in DMEM at 37°C and 10% CO_2_.

### Measurement of the contractile response of airways

Airway reactivity was assessed by measuring airway lumen changes using a phase-contrast microscope with video recording and solution perfusion capabilities, as previously described.(*17*) S1P (1 and 3 μM) and Sa1P (1 and 3 μM) were infused or added for incubation of the PCLS for 5 min, 10 min, or 15 hours.

### Measurements of intracellular Ca^2+^

Intracellular Ca^2+^ oscillations in airway smooth muscle cells were visualized using a confocal microscope in PCLS prepared from transgenic mice expressing the GCaMP6 reporter as previously reported (PMID: 35776523). Fluorescence imaging was performed using a video-rate confocal microscope.(*43*) Changes in fluorescence intensity were analyzed by selecting regions of interest (ROI). Final fluorescence values (F) were expressed as a fluorescence ratio (F/F0) normalized to the initial fluorescence (F0).(*44*)

### Statistics

A 2-sample *t-*test or χ^2^ with Fisher’s exact test was used to compare 2 groups depending on the data type. One-way or 2-way ANOVA with the Šidák or the Tukey multiple comparisons test was used to perform univariable or multivariable association analyses. Nonparametric testing was performed where data were not normally distributed. To address multiple comparisons and control for the FDR, we applied the Benjamini-Hochberg (BH) procedure (32) to analyses involving 3 or more comparisons, using a maximum FDR of 0.05. Associations for SNPs and individual sphingolipids were analyzed using Pearson’s correlation coefficient (see Supplemental Methods for complete details). For all tests, differences were considered significant when *P* < 0.05, and the three significance levels are indicated as follows: \**P* < 0.05, \*\**P* < 0.01, and \*\*\**P* < 0.001. GraphPad Prism version 8.2 was used for all statistical analyses. A complete description of all methods used in this study is provided in this article’s supplement section.

### Study approval

The study was approved by the Weill Cornell IRB and New York Hospital Queens (protocol 19-08020637). All animal studies were conducted under protocols approved by the Institutional Animal Care and Use Committee of Weill Cornell Medicine.

## Supplementary Information

### Supplemental Methods

Supplemental Figures

Supplemental Table

## Data Availability

All data produced in the present study are available upon reasonable request to the authors

## Acknowledgments

We acknowledge the valuable statistical support provided by Zoe Verzani MPH, part of the Biostatistics Services at Weill Cornell Medicine.

## Author contribution

All authors contributed substantially to this study.

Conceptualization: AH, JPZ, and SW

Methodology: AH, JPZ, TW and SW

Investigation: AH, SB, JPZ, and SW

Funding acquisition: AH and SW

Analysis and interpreted the data: AH, JPZ, SW, and TW

Writing – original draft: AH, JPZ, TW, and SW

Writing – review & editing: AH, JPZ, TW, and SW

