## Supplemental for "Sphingosine-1-phosphate – sphinganine-1-phosphate Imbalance Drives Airway Hyperreactivity"

### Supplementary Material & Methods

#### Study Design/Subject Enrollment

Parents of children between the ages of 2 and 21 years were recruited for enrollment in this study with prevalent cases from three hospitals in New York City, NY, which provide pediatric primary care and pediatric pulmonology subspecialty care between May 2020 and May 2025. The hospitals are located in the boroughs of Manhattan, Brooklyn, and Queens and provide outpatient care. Case and Control subjects were recruited at the time of arrival at their pediatric appointment (routine pediatric primary care visit for controls, or asthma-specific pulmonology visits for cases) and were enrolled in the study. Case and Control subjects were enrolled concomitantly and sequentially over time if they met inclusion/exclusion criteria (described below) without consideration of gender or age.

Children were classified as having asthma if they were given a diagnosis of asthma by the pediatric pulmonologist with whom they were an established patient and had been symptomatic within the last 2 years. Children with asthma were enrolled in the study at the time of their visit with the pulmonologist. Healthy control subjects were recruited out of the general pediatric practices at routine annual visits and were provided a screening questionnaire to evaluate for any prior history of wheezing, bronchodilator administration by a provider or parent, or any history of breathing concerns. An affirmative history of difficulty breathing in any context or use of bronchodilator led to the exclusion of the subject. Subjects for both case or control groups were additionally excluded if they reported being pregnant or if they have any additional diagnoses (e.g., HIV, sepsis, etc.) that may alter immune or inflammatory responses that are not related to the diagnoses relevant to this study. The study was approved by Weill Cornell, New York Hospital Queens, and Brooklyn Institutional Review Board (protocol #19-08020637) for human subjects. Following informed consent, all subjects underwent collection of peripheral blood for whole blood sphingolipid quantification, DNA isolation, and genotyping at the 17q21-associated SNP.

**Quantitative Sphingolipid Determination**

Sphingolipids were quantified in the whole blood of children and murine dry blood spots by high-pressure liquid chromatography electrospray ionization tandem mass spectrometry (HPLC-MS/MS) using a minor modification of a described method.[1] The method is validated for 5 dihydroceramides: (d18:0/16:0 d18:0/18:0, d18:0/18:1, d18:0/24:0, d18:0/24:1), 6 ceramides (d18:1/C16:0, d18:1/C18:0, d18:1/C20:0, d18:1/C22:0, d18:1/C24:0, d18:1/C24:1), 4 sphingomyelins (SM d18:1/C16:0, SM d18:1/C18:0, SM d18:1/C18:1, SM d18:1/C24:1), and 4 long-chain bases: sphingosine (SO d18:1), sphinganine (SA d18:0), sphingosine-1-phosphate (S1P d18:1), sphinganine-1-phosphate (Sa-1-P d18:0). 25 ul whole blood were extracted by vortexing overnight in 900 ul dichloromethane / methanol (1:1) with addition of internal standard (N-lauroyl-D-erythro-sphingosylphosphorylcholine). After centrifugation to precipitate cell debris, an aliquot was transferred into an Agilent 1200 HPLC (Agilent Poroshell 120 column) linked to an Agilent 6430 triple quadrupole mass spectrometer. Mobile phase A consisted of methanol/water/chloroform/formic acid (55:40:5:0.4 v/v); Mobile phase B consisted of methanol/acetonitrile/chloroform/formic acid (48:48:4:0.4 v/v). After pre-equilibration for 6 sec, the gradient was increased gradually to 60% mobile phase B and 100% mobile phase B that was held for 1.9 min. With a flow rate is 0.6 mL/min, the duration of the entire run was 9.65 min. We used the Mass Hunter optimizer and pure synthetic standards (Avanti Polar Lipids) to determine optimum fragmentation voltage, precursor/ product ions, and m/z values. Peak calls and abundance calculations were obtained with MassHunter Workstation Software Version B.06.00 SP01/Build 6.0.388.1 (Agilent). Final concentrations were calculated from a standard curve for each sphingolipid run in parallel.

**SNP genotyping**

DNA was isolated, and genotyping for the rs7216389 SNPs was performed on whole blood from both asthma and control subjects. Briefly, genomic DNA was extracted from 200µl whole blood using QIAamp DNA blood mini kit (QIAGEN Inc., cat# 51106) according to manufacturer's instructions, and the concentration quantitated by UV absorbance. The SNP genotyping was performed using the TaqMan® SNP Genotyping Assays(SNP ID: rs7216389) following the manufacturer's instructions. Each SNP genotyping reaction was carried out in duplicate, and three positive controls were included for the SNP genotype. The thermal cycling conditions included an initial incubation at 50°C for 2 min, then 95°C for 10 min, and 40 cycles of 95°C for 15 seconds and 60°C for one minute. The SNP genotyping reaction was run in an Illumina Eco Real-Time PCR system and the data was analyzed using EcoStudy (version 5.0.4890) software.

**Reagents**

Sphingosine-1-phosphate (S1P) was solubilized in 100% methanol, heated to 65°C, and sonicated until completely dissolved. The solution was then aliquoted into glass vials (2 mM stock solution). The S1P stocks were stored at −20°C until needed. On the day of the experiment, methanol was evaporated by slowly rotating the vials in a warm water bath (approximately 50°C) until a thin film of S1P was visible. Fatty acid-free bovine serum albumin was added (BSA, 4mg/ml) in Hank's balanced salt solution (HBSS) supplemented with 20 mM HEPES buffer and adjusted to pH 7.40 (sHBSS) at 37°C to a glass vessel containing S1P to make a 0.1, 0.3, 1, 3, or 10uM. The final solutions were incubated for 30 minutes and occasionally vortexed.

Sphinganine-1-phosphate (Sa1P) was dissolved in dimethyl sulfoxide (DMSO) to prepare the 3 mM stock solution; on the day of the experiment, the stock solution was diluted in sHBSS to prepare a working solution.

JTE-013, an S1P receptor two antagonist ($S1P_2$), and CAY10444 S1P receptor three antagonists ($S1P_3$) were solubilized in DMSO to prepare 10 mM stocks and subsequently diluted in sHBSS to achieve a final concentration of 10 μM.

**Mouse studies**.

All animal studies were conducted under protocols approved by the Weill Cornell Medical College Institutional Animal Care and Use Committee. Female C57Bl/6 mice, obtained from Taconic Farms, were housed under specific pathogen-free conditions and used at 10 to 14 weeks of age.

Heterozygous SPT-deficient mice (Sptlc2$^{+/-}$) or homozygous controls (Sptlc2$^{+/+}$), originally provided by Xian-Cheng Jiang, State University of New York (SUNY) Downstate Medical Center, were bred, identified by genotyping,[2] and used at 10-14 weeks of age.

**Evaluation of small airway contractility in precision cut lung slices (PCLS)**

Mice were euthanized by intraperitoneal injection pentobarbital (100 mg/kg). The trachea was cannulated with an intravenous catheter tube (20G Intima; Becton Dickinson), connected to a catheter extension and a 4-way stopcock. A syringe filled with 3 ml of air was attached to one port while the other port was closed. The chest cavity was opened by cutting along the sternum and the ribs adjacent to the diaphragm. An agarose solution was made by dissolving and melting 0.2g of low gelling temperature agarose in 5 ml of de-ionized $H_2O$ heated at 95ºC and subsequently adding 5ml of 2x Hanks' balanced salt solution (HBSS) supplemented with 20 mM HEPES buffer and adjusted to pH 7.40 (sHBSS). The solution was kept at 39ºC until used for infusion. A syringe filled with the warm agarose solution (2% w/v in sHBSS) was attached to the second port of the catheter. The IV tube was clamped proximal to the trachea and purged of air with the agarose solution by allowing the trapped air to escape via a 27G needle inserted into the IV tube proximal to the clamp. The IV clamp was removed, and the lungs were re-inflated by injecting ~1.3 ml of the agarose solution. Subsequently, ~0.2 ml of air was injected into the airways to flush the agarose out of the airways and into the distal alveolar space. Immediately after agarose inflation, the lungs were soaked with ice-cold sHBSS, and the body of the animal was placed at 4°C for 15 min. The lung and heart were removed and placed in sHBSS (4°C) and cooled for an additional 30 min to ensure the complete gelling of the agarose.

To prepare PCLS, a single lung lobe was removed from the respiratory tree by cutting the main bronchus. The lung lobe was trimmed near the bronchus to produce a flat surface that adhered to the mounting block of a VF-300 vibratome (Precisionary Instruments, Greenville, NC) following manufacturer instructions. Serial lung slices (~130 µm thick) were collected in sHBSS maintained at 4°C, transferred to Petri dishes containing cell culture medium i.e., low glucose DMEM supplemented with 3.7 g/L $NaHCO_3$ and an antibiotic-antimytotic mixture (ThermoFisher), and then transferred to a cell culture incubator maintained at 37°C in a humidified environment with 10% $CO_2$. Long-term treatment of PCLS with Sphingosine-1-phosphate (S1P), sphinganine-1-phosphate (Sa1P) or DMSO was performed by incubating the freshly prepared PCLS for 15 hours in culture medium supplemented with the shpingolipids (or DMSO for controls) diluted at their final concentrations. After overnight incubation, the PCLS were removed from the cell culture incubator and transferred to sHBSS.

PLCS containing airways that completely attached to the surrounding parenchyma and with ciliary activity were selected and mounted into a custom-made perfusion chamber consisting of a Plexiglas support for a 22 × 40 mm cover glass. A PCLS was placed in the center of the cover glass and held in place by placing a nylon mesh with a small hole in the center on the top of the PCLS. This small hole was centered on the airway of interest to allow imaging and recording of contractile activity without the interference of the mesh. A second 11 × 22 mm coverglass edged with silicone grease (Valve sealant; Dow Corning Co.) was placed over the slice and nylon mesh. We selected airways with an average cross-section size of 150-300 µm and confirmed that there was no significant difference in basal (un-stimulated) airway size among the testing groups ($p > 0.05$, t-test).

Perfusion of the PCLS in the chamber was performed by applying a gravity-fed flow of solution at one end of the glass chamber and suction at the other end of the chamber. The solutions perfusion was controlled by using a custom-built perfusion system consisting of eight solution reservoirs (30 ml) connected to a manifold with a single output tube (Warner Instruments Inc.). The flow from each reservoir was regulated by a valve (LFVA; Lee Company) under TTL control generated by a custom-made electronic control interfaced with the image acquisition software (Video Savant, IO Industries). The lung slices were observed on an inverted microscope (Diaphot 300; Nikon) with a 20× objective. Images were recorded with a CCD camera using a frame grabber PC card (Picolo; Eurosys) and the image acquisition software (Video Savant; IO industries, Inc.). Digital images (648 × 484 pixels) were recorded in time-lapse (0.5 Hz) and stored directly on a hard drive. The cross-sectional lumen area of the airway in each image was obtained by pixel summing using a custom-made software that runs in Video Savant, while the images are being captured. The airway lumen area was normalized to the initial area, i.e. before the perfusion of any agonist or drug, and the changes in lumen area over time were plotted.

To evaluate airway hyperreactivity, PCLS were initially infused with sHBSS for 3 min, followed by infusion of S1P and/or Sa1P diluted in sHBSS at the concentrations indicated in the figures and legends and for the duration also indicated in the figures.. Solution changes in the perfusion chamber were performed according to a pre-programmed time schedule and using a customized and computer-controlled perfusion system coupled to the imaging capture and analysis setup. Methacholine (MCh) was dissolved in de-ionized $H_2O$ to make a stock solution and dissolved in the sHBSS to its final concentration on the same day of the experiments.

**Measurements of intracellular $Ca^{2+}$**

PCLS were prepared from transgenic mice expressing the GCaMP6 calcium reporter in all cells and tissues. Lung slices were mounted in the perfusion chamber as described for the evaluation of airway contractility. Fluorescence imaging was performed using a video-rate confocal microscope.[3] A 488-nm laser supplied the excitation wavelength, and the resultant fluorescence images (>510 nm) were recorded at 15 Hz. Changes in fluorescence intensity were analyzed by selecting regions of interest (ROI) ranging from 5 to 7 pixels[2]. The average fluorescence intensities of an ROI were obtained, frame by frame, using custom-written software that allowed the tracking of the ROI within an SMC as it moved with contraction. Final fluorescence values (F) were expressed as a fluorescence ratio (F/F0) normalized to the initial fluorescence (F0). Line scan analysis of images was performed by extracting a line of pixels from each image and placing them sequentially to form a time sequence in a single image.[4]

**Supplemental Figures and Figure Legends**

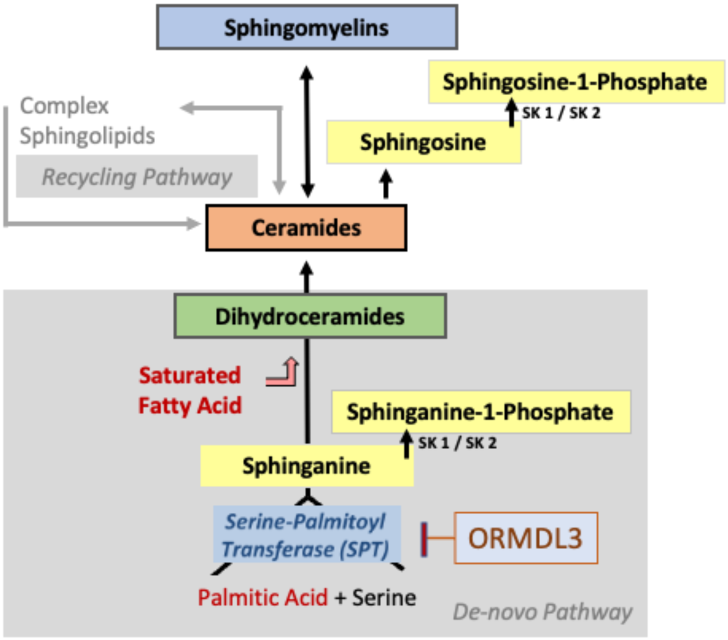


**Supplemental Figure 1. Sphingolipid synthesis pathways.** ORMDL3 is a regulator of sphingolipid synthesis which encodes transmembrane proteins localized in the endoplasmic reticulum, where it acts to negatively regulate sphingolipid synthesis through inhibition of serine palmitoyl-CoA transferase (SPT) which is the rate-limiting enzyme in *de novo* sphingolipid synthesis. Sphingosine kinases (SK) 1 and 2 generate sphingosine 1-phosphate (S1P) from sphingosine, and sphinganine 1-phosphate (Sa-1P) from sphinganine.

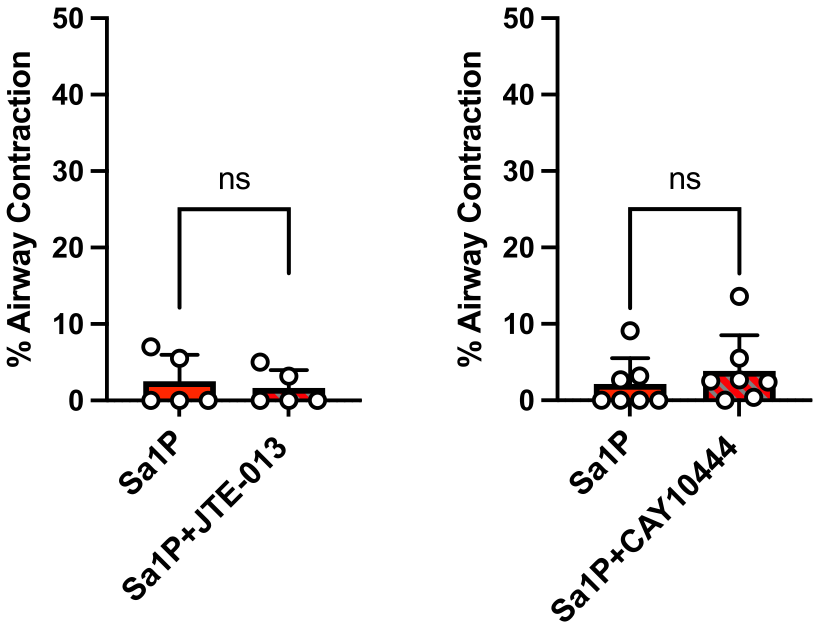


**Supplemental Figure 2. The effect of S1P receptor antagonists on airway contraction stimulated by Sa1P.** Summary data showing the effect of S1P receptor 2 antagonist JTE-013 (10 µM ) and S1P receptor 3 antagonist CAY10444 (10 µM) on the Sa1P, (**A and B**) induced airway contraction. Data are means ± SEMs of 5– 10 airways from four mice in each group. *$p < 0.05$, unpaired *t*-test.

| | **Overall**<br>N = 342[1] | **Asthma**<br>N = 232[1] | **Control**<br>N = 110[1] | **p-value**[2] |
|---|---|---|---|---|
| Genotype | | | | <0.001 |
| CC | 31 (9.1%) | 13 (5.6%) | 18 (16%) | |
| CT | 129 (38%) | 73 (31%) | 56 (51%) | |
| TT | 182 (53%) | 146 (63%) | 36 (33%) | |
| Asthma Severity | | | | >0.9 |
| Intermittent Asthma | 69 (31%) | 69 (31%) | 0 (NA%) | |
| Mild Persistent Asthma | 66 (29%) | 66 (29%) | 0 (NA%) | |
| Moderate Persistent Asthma | 72 (32%) | 72 (32%) | 0 (NA%) | |
| Severe Persistent Asthma | 19 (8.4%) | 19 (8.4%) | 0 (NA%) | |
| No Asthma | 116 | 0 | 116 | |
| Gender | | | | 0.2 |
| Female | 170 (50%) | 121 (53%) | 49 (45%) | |
| Male | 169 (50%) | 109 (47%) | 60 (55%) | |
| Age | | | | 0.5 |
| Mean (SD) | 10.9 (5.7) | 10.8 (5.3) | 11.2 (6.4) | |
| Median (Q1, Q3) | 11.5 (6.0, 16.0) | 10.0 (6.0, 15.0) | 14.0 (6.0, 17.0) | |
| Min, Max | 0.0, 21.0 | 1.0, 21.0 | 0.0, 21.0 | |
| Ethnicity | | | | 0.033 |
| Hispanic or Latino | 178 (53%) | 129 (57%) | 49 (45%) | |
| Not Hispanic or Latino | 156 (47%) | 96 (43%) | 60 (55%) | |
| Unknown | 8 | 7 | 1 | |
| Race | | | | <0.001 |

| | Overall<br>N = 342[1] | Asthma<br>N = 232[1] | Control<br>N = 110[1] | p-value[2] |
|---|---|---|---|---|
| Asian | 36 (15%) | 18 (11%) | 18 (22%) | |
| Black | 100 (41%) | 77 (48%) | 23 (28%) | |
| Other | 27 (11%) | 22 (14%) | 5 (6.0%) | |
| White | 81 (33%) | 44 (27%) | 37 (45%) | |
| Unknown | 98 | 71 | 27 | |

[1] n (%)
[2] Pearson's Chi-squared test; Fisher's exact test; Welch Two Sample t-test

**Supplemental Table 1. Genotype distribution and demographic characteristics of the children's cohort.** The table summarizes genotype frequencies (CC, CT, TT) and demographic variables (asthma severity, sex, age, ethnicity, and race) among children with asthma (n = 232) and controls (n = 110). Enrichment of the TT genotype was observed in the asthma group ($p < 0.001$). No significant differences were found in asthma severity, sex, or age between groups. Ethnicity differed modestly ($p = 0.033$), while race showed a significant distribution difference ($p < 0.001$). Statistical comparisons were performed using Pearson's Chi-squared test, Fisher's exact test, or Welch's t-test as appropriate.

**Graphical Abstract**

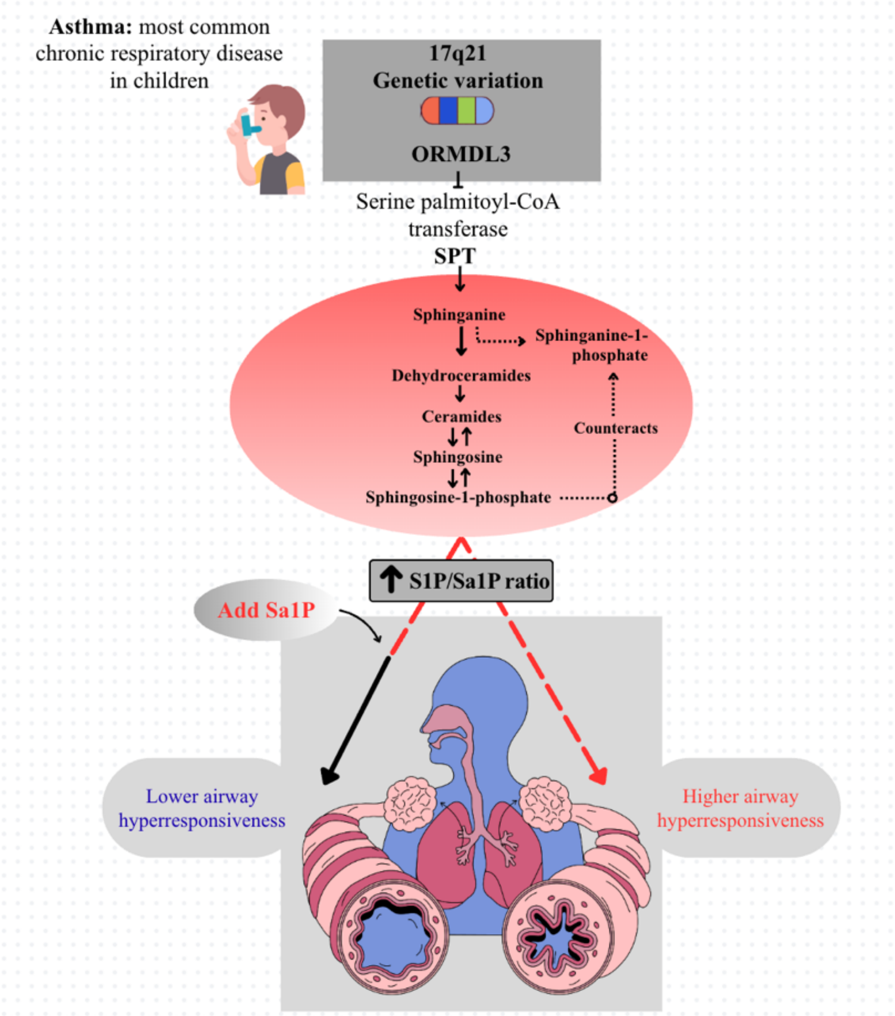
Asthma: most common chronic respiratory disease in children
17q21
Genetic variation
ORMDL3
Serine palmitoyl-CoA transferase
SPT
Sphinganine
Sphinganine-1-phosphate
Dehydroceramides
Ceramides
Sphingosine
Sphingosine-1-phosphate
Counteracts
S1P/Sa1P ratio
Add Sa1P
Lower airway hyperresponsiveness
Higher airway hyperresponsiveness
